# Dev’s Formulae for Diagnostic Test Accuracy Meta-analysis

**DOI:** 10.1101/2023.03.27.23287186

**Authors:** Dev Desai

**Affiliations:** Smt. NHLMMC, Ahmedabad

**Keywords:** Sensitivity, Specificity, PPV, True Positive, False Positive, True Negative, False Negative

## Abstract

Calculating the sensitivity and specificity values from True Positive, True Negative, False Positive, and False Negative is easy. But estimating the True and False values back is not possible. All the formulae present do that need the value of the disease’s prevalence in the population and hospital-based studies are usually biased in the prevalence. We have tried to create formulae and have been successful along with its proof on formulae that uses Sensitivity, Specificity, PPV and Total participants of the study to calculate True and False values.

---

Testing for Diagnostic accuracy of different scores and different methods has become increasingly vital as the world progresses towards Evidence based Medicine. Its importance is reflected by the fact that all new treatment plans are developed and are tested against a gold standard treatment plan to see the prognosis as well as its feasibility.

It is not new to test any new technique for diagnosis against a gold standard and comparing the results to see the sensitivity and specificity of the new test along with the diagnostic accuracy of test in order to decide whether the new test is usable or not. Various papers in many different fields and clinical settings have been published, which have already tested various methods, thereby either prove their worth or reject them conclusively.

The problem arises at the time of secondary analysis of the data as most publications and journals publish the data analysis but do not publish raw data. In papers with published data on diagnostic test accuracy, the paper would contain the values of Sensitivity and specificity and the total patients enrolled, but it doesn’t contain the actual values of how many patients actually had the disease and out of which, how many were diagnosed perfectly from the new technique. This value is called the True Positive (TP).

It is impossible to calculate values of True Positive (TP), False Negative (FN), True negative (TN), and False Positive (FV).

The same problem was encountered while doing a Diagnostic accuracy test between different scoring methods. Due to this, there was a need to find the original values and hence, the below presented formulae were derived mathematically. Hereby, Using the Values of Sensitivity, Specificity, PPV, and Total Participants, it is possible to calculate TP, FP, FN, and TN.

|  | Disease Present | Disease Absent |  |
| --- | --- | --- | --- |
| Test Positive | TP (A) | FP (D) | A + D |
| Test Negative | FN (B) | TN (C) | B + C |
| | A + B | C + D | $x = A + B + C + D$ |

TP (A) = True Positive (denoted as “A” in the equations)
FN (B) = False Negative (denoted as “B” in the equations)
TN (C) = True Negative (denoted as “C” in the equations)
FP (D) = False Positive (denoted as “D” in the equations)
Total (X) = TP + FN + TN + FP = A + B + C + D
Sensitivity = Sen
Specificity = Spe
PPV = Positive Predicted Value

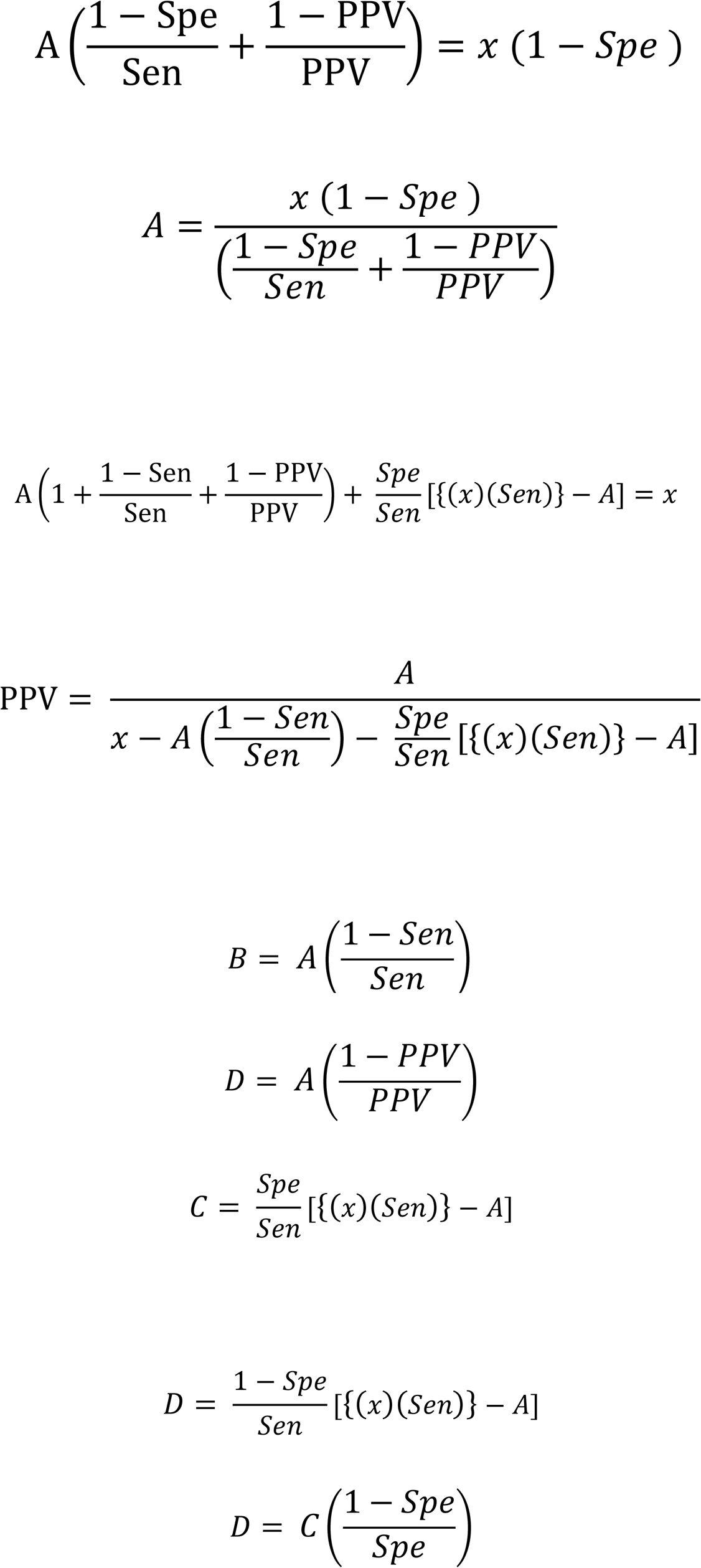

To test the validity of above formulae, a pool of 5 different Diagnostic test accuracy research results was identified by selecting sequential papers which had given the values of Sensitivity, Specificity, PPV, and total participants, along with true values of patient identification using their diagnostic tool. Its Bland – Altman Analysis was carried out.

**Table 1.**
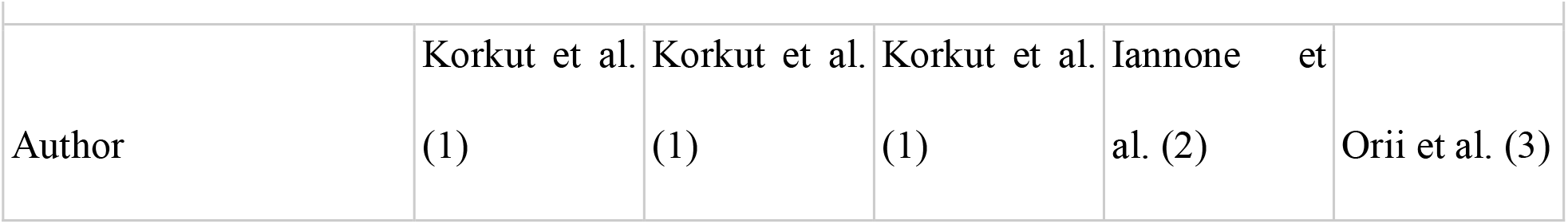
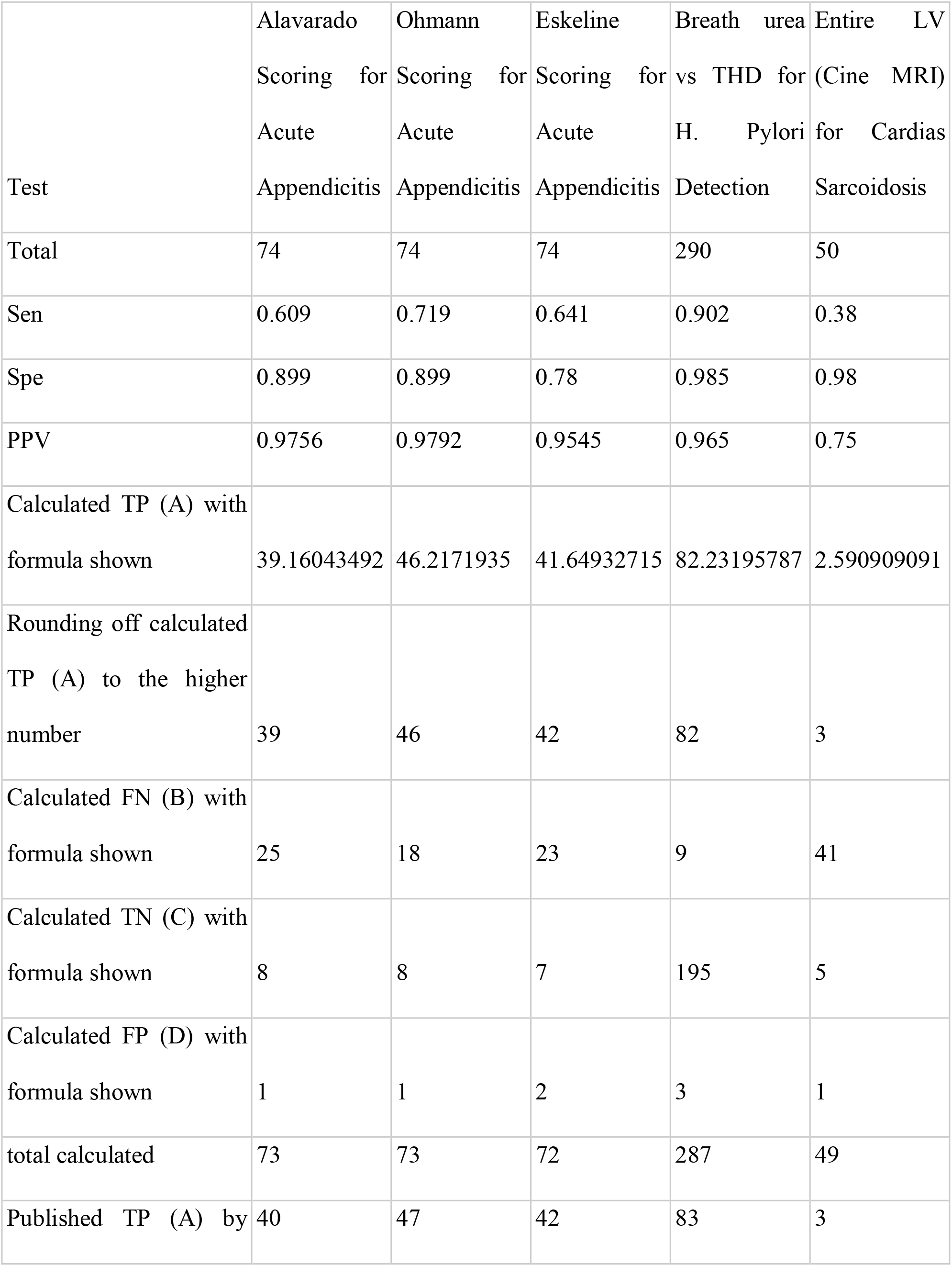
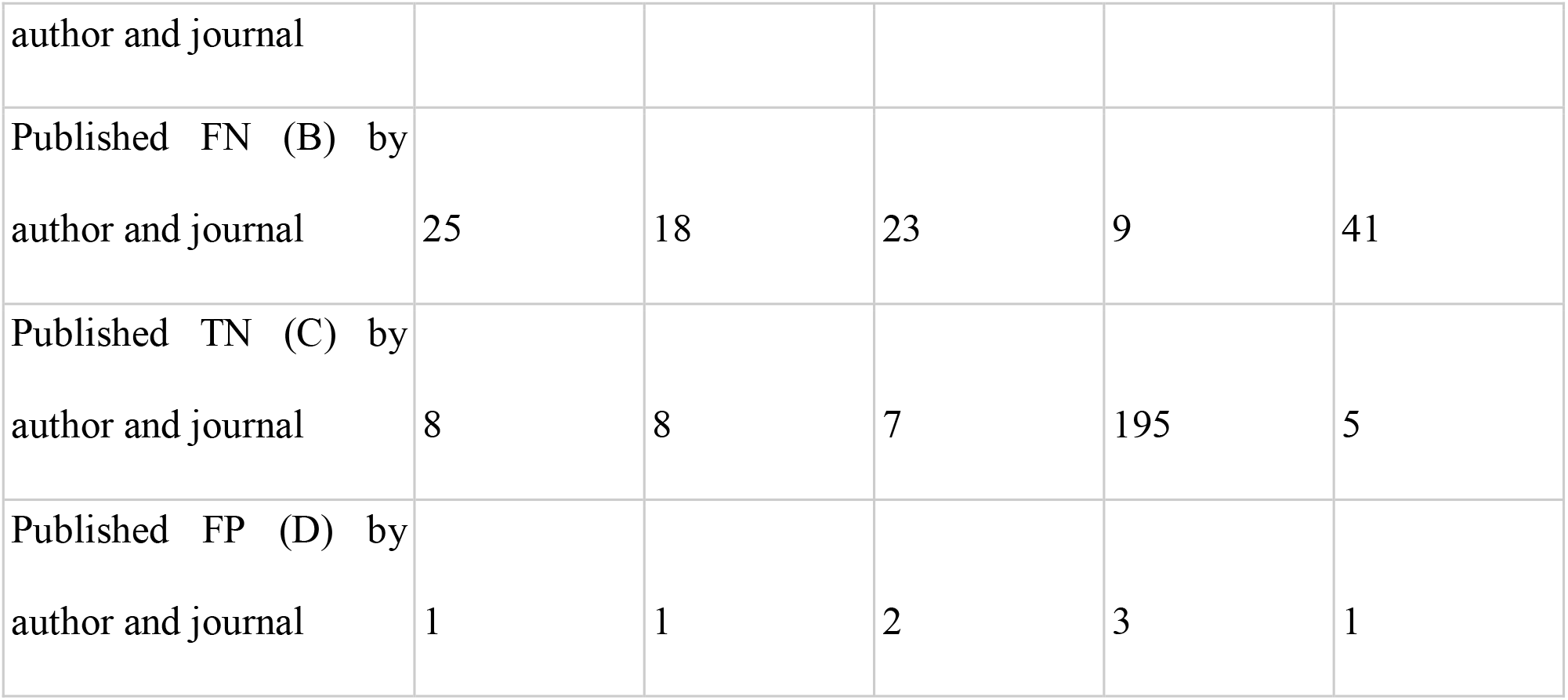
5 different diagnostic accuracy tests results for validation of the formulae

The table above shows that when the calculated TP(A) is rounded off to the closest higher number, it is identical to the actual value of True Positive patients published by the author and the journal. The Rounded off value can be used further to calculate the values of FN(B), TN(C), and FP(D) using the formulae shown above and their total is identical to the total participants taken by the author.

Rounding off to higher number might produce a problem that total number of patients calculated with the formulae might be higher than the patients enrolled but the sensitivity, specificity remains exactly the same of the calculated data and this ensures that in the secondary analysis when more papers would be added, there will not be any bias created.

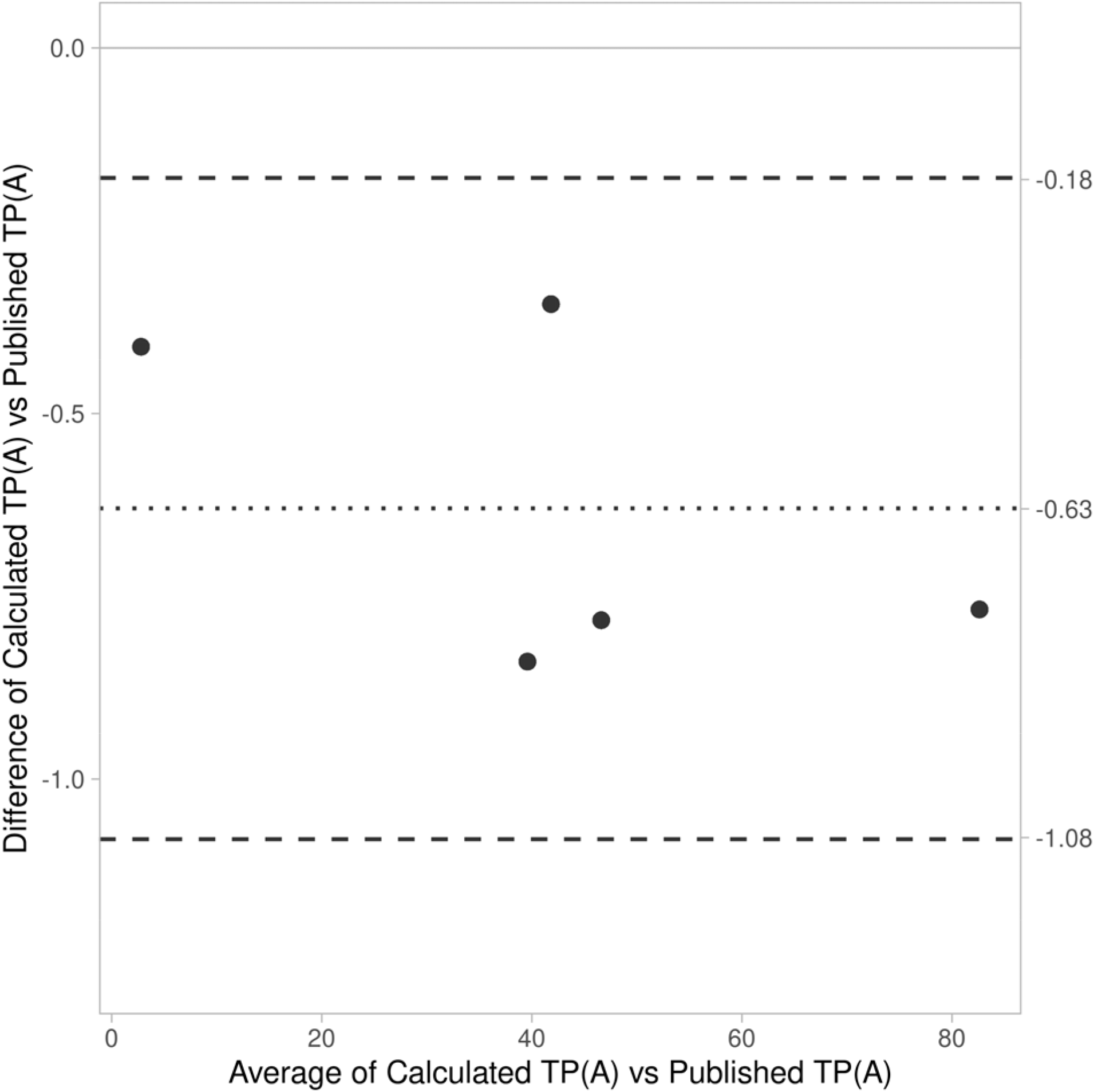

The Bland Altman analysis here gives the Mean difference of -0.63 with 95% CI ranging from -1.08 to -0.18, being Statistical Significant.

This shows that the formulae created can be used successfully to find out the True values from analyzed data (where raw data is unavailable) and different combinations and derivatives of these formulae can prove an important statistical tool to authors doing a review or a meta or secondary analysis of already published data. This formula can be used by software to directly calculate the True Values reducing the author’s workload of gathering the true values and makes analysis easier, especially in the cases of Diagnostic Test Accuracy Analysis.

## Data Availability

All data produced in the present work are contained in the manuscript

## Notes

### Competing Interest Statement

The authors have declared no competing interest.

### Funding Statement

This study did not receive any funding

